# Diabetes-related excess mortality in Mexico: a comparative analysis of national death registries between 2017-2019 and 2020

**DOI:** 10.1101/2022.02.24.22271337

**Authors:** Omar Yaxmehen Bello-Chavolla, Neftali Eduardo Antonio-Villa, Carlos A. Fermín-Martínez, Luisa Fernández-Chirino, Arsenio Vargas-Vázquez, Daniel Ramírez-García, Martín Roberto Basile-Alvarez, Ana Elena Hoyos-Lázaro, Rodrigo M. Carrillo-Larco, Deborah J. Wexler, Jennifer Manne-Goehler, Jacqueline A. Seiglie

## Abstract

**BACKGROUND:** Excess all-cause mortality rates in Mexico in 2020 during the COVID-19 pandemic were among the highest globally. Recent reports suggest that diabetes-related deaths were also higher, but the contribution of diabetes as a cause of excess mortality in Mexico during 2020 compared to prior years has not yet been characterized.

**METHODS:** We conducted a retrospective, state-level study using national death registries from Mexican adults ≥20 years for the 2017-2020 period. Diabetes-related deaths were classified using ICD-10 codes that listed diabetes as the primary cause of death, excluding certificates which listed COVID-19 as a cause of death. Excess mortality was estimated as the increase in diabetes-related mortality in 2020 compared to average rates in 2017-2019. Analyses were stratified by diabetes type, diabetes-related complication, and in-hospital vs. out-of-hospital death. We evaluated the geographic distribution of diabetes-related excess mortality and its socio-demographic and epidemiologic correlates using spatial analyses and negative binomial regression models.

**RESULTS:** We identified 148,437 diabetes-related deaths in 2020 (177/100,000 inhabitants), 41.6% higher than the average for 2017-2019, with the excess occurring after the onset of the COVID-19 pandemic. In-hospital diabetes-related deaths decreased by 17.8% in 2020 compared to 2017-2019, whereas out-of-hospital deaths increased by 89.4%. Most deaths were attributable to type 2 diabetes and type 1 diabetes (129.7 and 4.0/100,000 population). Diabetes-related emergencies as contributing causes of death also increased in 2020 compared to 2017-2019 for hyperglycemic hyperosmolar state (128%), and ketoacidosis (116%). Diabetes-related excess mortality clustered in southern Mexico and was highest in states with higher social lag, higher rates of COVID-19 hospitalization, and higher prevalence of HbA1c ≥7.5%.

**INTERPRETATION:** Diabetes-related mortality increased among Mexican adults by 41.6% in 2020 after the onset of the pandemic compared to 2017-2019, largely attributable to type 2 diabetes. Excess diabetes-related deaths occurred disproportionately out-of-hospital, clustered in southern Mexico, and were associated with higher state-level marginalization, rates of COVID-19 hospitalizations, and higher prevalence of suboptimal glycemic control. Urgent policies to mitigate mortality due to diabetes in Mexico are needed, particularly given the ongoing challenges in caring for people with diabetes posed by the COVID-19 pandemic.

**Research in context:** *Evidence before this study:* We searched PubMed and Google Scholar for research articles published up to February 15, 2022, using the terms [(“diabetes-related mortality” OR (“excess mortality” AND “diabetes”))]. No language restriction was applied. This search revealed few international studies evaluating nationwide diabetes-related mortality in general. In Mexico, only one unpublished study evaluated diabetes-related mortality up to 2019. We identified no studies which evaluated diabetes-related excess mortality in Mexico or elsewhere during 2020 or which explored correlates of diabetes-related excess mortality in 2020.

*Added value of this study:* This is the first report and characterization of an excess in diabetes-related mortality in Mexico during 2020 compared to recent years. Diabetes as a primary cause of death in Mexico was higher in 2020 compared to 2017-2019, particularly for people living with type 2 diabetes, starting in March 2020 with the onset of the COVID-19 pandemic. Compared to the 2017-2019 period, most of these excess deaths occurred out of hospital, with a concurrent decrease in in-hospital diabetes-related mortality. Hyperosmolar hyperglycemic state and ketoacidosis as primary causes of diabetes-related deaths also increased in 2020 compared to prior years. Our study also identified substantial geographic variation in diabetes-related excess mortality in Mexico, with southern, poorer States bearing a disproportionate burden. Finally, we report that diabetes-related excess mortality was associated with higher marginalization, suboptimal glycemic control, and higher rates of COVID-19 hospitalization, which were clustered in southern Mexico.

*Implications of the available evidence:* Readily treatable, high morbidity diabetes-related conditions were likely untreated due to the constraints of the health care system during the COVID-19 pandemic, leading to diabetes-related excess mortality. This is a problem for Mexico, but it is likely to be generalizable to other countries and other conditions, as seen even in high-income countries. Given the ongoing challenges posed by the COVID-19 pandemic on healthcare systems, policies that can strengthen care for diabetes and other chronic conditions are urgently needed to mitigate the dramatic rise in diabetes-related mortality occurring in the out-of-hospital setting and its disproportionate burden on populations with high levels of marginalization.

## INTRODUCTION

Diabetes mellitus is a leading cause of disability, morbidity, and mortality world-wide. In Mexico, diabetes is the second leading cause of death and has an estimated prevalence of 15.2% (12.8 million adults)^1,2^; furthermore, over the past three decades, mortality attributable to diabetes increased by an alarming 77%^3^. In 2020, during the COVID-19 pandemic, Mexico experienced one of the highest rates of all-cause excess mortality globally (46.5% increase from prior years). Even though the majority of these excess deaths were directly attributable to COVID-19 case mortality, reports by the National Institute for Statistics and Geography (INEGI) suggest that excess deaths in Mexico during 2020 were also attributable to an increase in mortality from non-COVID-19 causes, including cardiovascular disease and diabetes^4^. However, the extent to which diabetes contributed as a cause of this excess mortality during 2020 compared to recent years has not yet been characterized^5^.

Diabetes prevalence, as well as diabetes-related complications and mortality, are tightly associated with socio-demographic inequalities^2,3^. These inequalities were unmasked by the COVID-19 pandemic, in part due to a fragmented care infrastructure that preceded COVID-19 and interruptions in in-person care, which disproportionately impacted socioeconomically vulnerable populations^6,7^. As such, while the intersection of diabetes and COVID-19 and their compounded severity was an important contributor to all-cause excess mortality in Mexico in 2020^8–10^, the pandemic had ripple effects that also impacted care continuity for people with diabetes independent of COVID-19 itself. Furthermore, because hospital saturation was highest in marginalized communities^8,11,12^, we hypothesized that diabetes-related deaths were higher in 2020 compared to prior years and that these excess deaths were associated with higher levels of marginalization. Given that the COVID-19 pandemic is ongoing and continues to pose a significant burden on health systems globally, characterizing the extent to which diabetes-related mortality rates may have increased in 2020 could help guide policies to mitigate interruptions in diabetes care and strengthen existing systems for diabetes care delivery across the healthcare system.

In this study, we sought to characterize: 1) the age-adjusted rates of diabetes-related excess mortality among Mexican adults ≥20 years during 2020, overall and stratified by diabetes type, preventable diabetes emergencies and complications as contributing causes of death, and in-hospital vs. out-of-hospital deaths; 2) the geographic distribution of diabetes-related excess mortality in Mexico; and 3) socio-demographic and epidemiologic correlates of diabetes-related excess mortality in Mexico in 2020.

## METHODS

### Study design and data source

We conducted a retrospective, state-level study using national death registries obtained from the dynamic information cubes registered by INEGI for the 2017-2020 period, last updated on October 21^st^, 2021. Briefly, INEGI generates annual mortality statistics from death certificates issued by the Ministry of Health. Death registries are comprised of systematic daily mortality records, which are coded using the tenth revision of the International Classification of Diseases (ICD-10).

### Variables and definitions

#### Outcome variables

Our analysis was centered on two primary outcomes: Diabetes-related mortality and diabetes-related excess mortality.

1. *Diabetes-related mortality* – Defined according to death certificates generated by INEGI, which listed one of the following ICD-10 codes as the primary cause of death: E10-Type 1 diabetes mellitus, E11-Type 2 diabetes mellitus, and E12-14-Other diabetes mellitus (including malnutrition-related diabetes, other specified diabetes mellitus, and unspecified diabetes mellitus). Diabetes-related mortality was additionally stratified by diabetes-related emergencies and complications listed as contributing causes in each death certificate and defined based on the following ICD-10 codes: hyperglycemic hyperosmolar state (HHS) (E10-14.0), ketoacidosis (E10-14.1), kidney complications (E10-14.2), ophthalmic complications (E10-14.3), neurological complications (E10-14.4), circulatory complications (E10-14.5), other specified and unspecified complications (E10-14.6 and E10-14.8), multiple complications (E10-14.7), and without complications (E10-14.9). To focus our analysis on diabetes-related mortality only, we excluded all causes of death codified as COVID-19 (ICD-10 code U07.1 or U07.2) during 2020. We also excluded deaths which did not have registered socio-demographic or geographic information. A flow diagram with study inclusion criteria is provided in **Supplementary Figure 1**.
2. *Diabetes-related excess mortality* – Estimated as the increase in diabetes-related mortality in 2020 compared to the 2017-2019 period average. This approach is consistent with the definition of excess mortality proposed by Karlinsky and Kobak, which allows for the comparison of all-cause excess mortality across countries and minimizes year to year variations in mortality^13^. Excess deaths were standardized to age-adjusted rates per 100,000 using population age structures by region and state per 5-year increments using population projections provided by the National Population Council^14^. Diabetes-related excess mortality was estimated at the regional level for descriptive purposes and at the state level for modeling (**Supplementary Material)**. Diabetes-related excess mortality is also presented as percent increase in 2020 compared to the 2017-2019 period.

#### In-hospital vs. out-of-hospital death

We hypothesized that the COVID-19 pandemic posed a significant burden on inpatient care, which may have influenced increases in age-adjusted diabetes-related excess mortality. To evaluate this, we stratified diabetes-related mortality by whether the death occurred in-hospital vs. out-of-hospital, as registered in death certificates. Out-of-hospital deaths were defined accordingly if the death was not registered in a hospital setting or if they were coded as occurring at the deceased’s home or elsewhere (i.e., in the streets in some instances). Deaths without specified place of death were classified as unspecified and were excluded from these analyses (2274 deaths in 2020, 1433 in 2019, 1187 in 2018, and 1206 in 2017). Next, we calculated the ratio of the number of deaths which occurred out-of-hospital divided by those which occurred in-hospital per state and year. We considered the differences in this ratio for 2020 compared to the average ratio in 2017-2019 as the main measure for this outcome.

#### Epidemiologic indicators of diabetes care and diabetes prevalence

To evaluate the association between diabetes-related excess mortality and epidemiologic indicators with relevance to diabetes care and diabetes prevalence in Mexico, we included three key state-level variables: 1) adequate glycemic control (defined as an HbA1c level ≥7.5%^15^), 2) undiagnosed diabetes prevalence, and 3) type 2 diabetes prevalence. These indicators were derived from the Mexican National Health and Nutrition Survey 2020 (ENSANUT COVID 2020)^16^. Prevalence estimates were computed using population weights from ENSANUT COVID 2020 with the *survey* R package.

#### Marginalization Index

To evaluate the association between marginalization and diabetes-related excess mortality, we used the population density Independent Social Lag Index (DISLI, further details in **Supplementary Materials**),^11,12^ which is a well-characterized state-level proxy of socio-demographic inequalities in Mexico.

#### State-level epidemiologic indicators of the COVID-19 pandemic

To evaluate the association between COVID-19 and diabetes-related excess mortality, we also included variables of relevance to COVID-19 epidemiology in Mexico. Namely, COVID-19 seroprevalence, COVID-19 hospitalization rates, incidence of COVID-19, and COVID-19 deaths in people with diabetes. These estimates were derived from the General Directorate of Epidemiology of the Mexican Ministry of Health dataset and ENSANUT COVID 2020 for COVID-19 seroprevalence data^17^.

### Statistical Analysis

First, to visualize differences in diabetes-related mortality in the 2017-2020 period, we plotted diabetes-related deaths per 100,000 inhabitants by month of occurrence, overall and stratified by diabetes type. We also plotted the overall number of diabetes deaths, stratified by diabetes type, age group, and out-of-hospital vs. in-hospital setting, and compared these estimates between the 2017-2019 average and 2020. We then disaggregated the diabetes-related mortality rates per 100,000 inhabitants by diabetes-related complications listed as contributing causes of death in the 2018-2020 period. The year 2017 was excluded due to incomplete ICD-10 codes for emergencies and complications related to diabetes-related deaths. This analysis was also stratified by out-of-hospital vs. in-hospital mortality.

Second, to visualize the geographical distribution of diabetes-related excess deaths in Mexico, we used chloropleth maps with the *ggmap* R package with the quantile method. To evaluate the spatial dependence of diabetes-related excess mortality and the out-of-hospital to in-hospital death ratio, we used Moran’s I statistic, which was obtained as an indicator of global spatial autocorrelation, and its significance was assessed through an inference technique based on randomly permuting the observed values over the spatial units. We also evaluated hotspots of excess mortality and the out-of-hospital to in-hospital death ratio to confirm autocorrelation using the Getis-Ordi Gi statistic.

Next, we used spatial autocorrelation to characterize whether key indicators relevant to diabetes care and diabetes prevalence in Mexico were associated with diabetes-related excess mortality. Similarly, we examined whether socio-demographic inequalities as proxied by DISLI, and COVID-19 indicators were associated with age-adjusted diabetes-related excess mortality. Bivariate correlations between epidemiological indicators and age-adjusted diabetes-related excess mortality were evaluated with Lee’s L test for spatial autocorrelation using spatial weights matrix within the *spdep* R package^18^. We also assessed the correlation of these epidemiological indicators with the out-of-hospital to in-hospital death ratio, as a proxy of their impact in access to medical care.

Finally, the simultaneous impact of all evaluated indicators on age-adjusted diabetes-related excess mortality was analyzed using negative binomial regression, with log-transformed population of each state as the regression offset. We used the Global Moran I test for regression residuals for all models and identified no significant spatial autocorrelation; therefore, we fitted all models without incorporating spatial effects. All statistical analyses were conducted using R software version 4.1.2

## RESULTS

### Diabetes-related mortality in Mexico in 2020 compared to 2017-2019

We analyzed data from 452,924 diabetes-related deaths in Mexico during the 2017-2020 period. We identified 148,437 diabetes-related deaths (177 per 100,000 inhabitants) in 2020, compared with an average of 101,496 deaths in 2017-2019 (125 per 100,000 inhabitants), which represents a 41.6% increase in diabetes-related deaths in 2020 compared to the average reported in the 2017-2019 period (**Figure 1**). Overall, the highest rate of diabetes-related mortality occurred after the month of May, with a peak during the June-July period after the onset of the COVID-19 pandemic (**Figure 1A**); notably these trends closely followed COVID-19 related mortality trends (**Supplementary Figure 2**). When stratified by diabetes type, the largest share of diabetes-related deaths in 2020 was attributable to type 2 diabetes (130 per 100,000 persons), and type 1 diabetes (3.99 per 100,000 persons). Compared with diabetes-related deaths in 2017-2019, these rates correspond to an excess mortality of 46.7% for type 2 diabetes and 53.5% for type 1 diabetes (**Figure 1B**). Diabetes-related mortality was higher than during the 2017-2019 period across all ages for all diabetes types (**Figure 1C**). Regarding place of death, out-of-hospital diabetes-related deaths increased by 89.4% from an average of 59,061 diabetes-related deaths (50.9 per 100,000 inhabitants) during 2017-2019 to 111,870 deaths (133.6 per 100,000 inhabitants) in 2020. Conversely, in-hospital deaths decreased by 17.8% from 41,176 diabetes-related deaths (50.9 per 100,000 inhabitants) in 2017-2019 to 33,825 deaths (40.4 per 100,000 inhabitants) in 2020. Overall, diabetes-related deaths shifted to the out-of-hospital setting by more than two-fold, as reflected by the increase in the out-of-hospital to in-hospital death ratio from an average of 1.43 in 2017-2019 to 3.31 in 2020. These increases were primarily attributed to an increase in out-of-hospital and a decrease of in-hospital deaths for type 2 diabetes. For type 1 diabetes, both out-of-hospital and in-hospital deaths increased (**Figure 1D**). Overall, the observed trends were similar for other diabetes types (**Supplementary Figure 3**).

**Figure 1.**
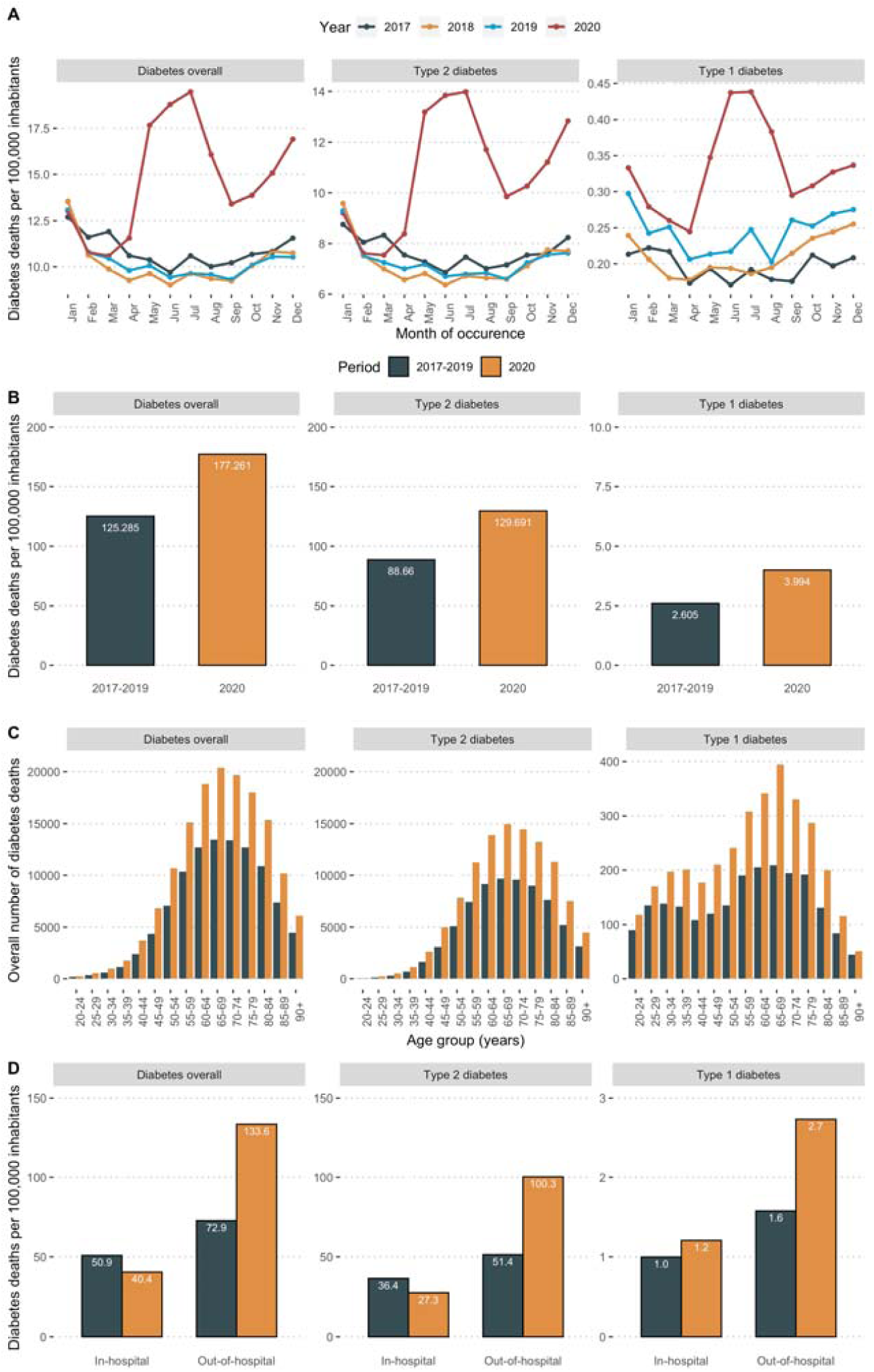
Monthly trends over time of diabetes-related deaths per 100,000 inhabitants during the 2017-2020 period and stratified for overall, type 1 and type 2 diabetes classified using ICD-10 criteria (A). We also show a comparison of diabetes-related deaths between the average of 2017-2019 and 2020 by diabetes type (B), by age group per 5-year increments (C) and stratified according to out-of-hospital vs. in-hospital mortality (C).

### Diabetes-related mortality in 2020 compared with 2017-2019, according to diabetes-related emergencies and complications as contributing causes of death

The stratification of diabetes-related mortality according to diabetes-related emergencies and complications as a contributing cause of death is presented in **Figure 2**. Compared with 2018-2019, the highest increase in diabetes-related complications as contributing causes of death in 2020 were HHS (128% absolute increase) and ketoacidosis (116% absolute increase). Other diabetes-related complications that increased compared to prior years were unspecified complications (69.5%), followed by kidney complications (24.9%), ophthalmic complications (22.2%), and lower-limb circulatory complications (15.1%). Notably, diabetes-related deaths with multiple complications and without complications also increased by 28.4% and 50.2%, respectively (**Figure 2A**). When stratified by place of death, we observed an increase in both in-hospital and out-of-hospital deaths for HHS (37.9% and 154% increase, respectively) and diabetic ketoacidosis (20.9% and 240% increase, respectively), while for kidney-related complications, we observed a 13.5% decrease in in-hospital and a 61.2% increase in out-of-hospital deaths (**Figure 2B**). Stratified analyzes that include all diabetes-related complications as contributing causes of death according to place of death are provided in **Supplementary Figure 4**.

**Figure 2.**
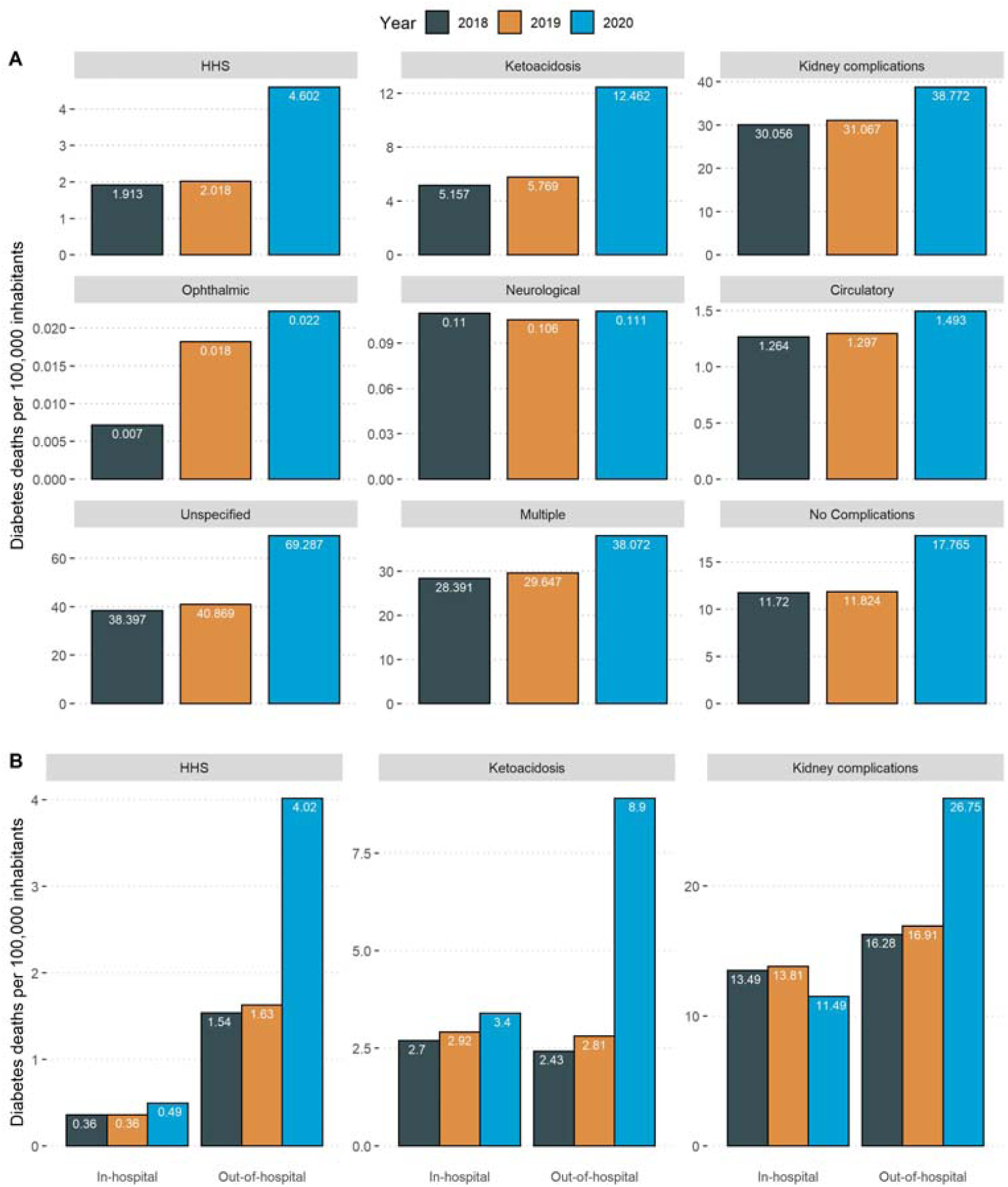
Diabetes-related mortality rates standardized per 100,000 population and disaggregated by diabetes-related emergencies and complications as contributing causes of death in 2018-2019 compared to 2020 (A). Contributing causes of death were classified using ICD-10 codes for each specific emergency and complication. The figure also shows the three leading diabetes-related emergencies and complications as contributing causes of death according to out-of-hospital vs. in-hospital mortality in 2020 compared to 2018-2019 (B). <u>**Abbreviations:**</u> HHS, Hyperglycemic Hyperosmolar State

### Geographic distribution and correlates of diabetes-related excess mortality in Mexico

We identified spatial autocorrelation in the geographic distribution of diabetes-related excess deaths in Mexico in 2020 compared to 2017-2019 (Moran’s I=0.335, p=0.002, **Figure 3A-C**). Diabetes-related excess mortality clustered near the Southeast and Gulf of Mexico regions, as identified using local indicators of spatial autocorrelation with Moran’s local statistic and the Getis-Ordi Gi statistic (**Supplementary Figure 5**). Similarly, the out-of-hospital to in-hospital mortality ratio increased markedly in 2020 compared to 2017-2019 and displayed a strong spatial autocorrelation which clustered within the same region (Moran’s I=0.481, p<0.001). This increase was primarily attributed to increased out-of-hospital diabetes-related mortality for type 2, but not for type 1 diabetes (**Supplementary Figure 6**).

**Figure 3.**
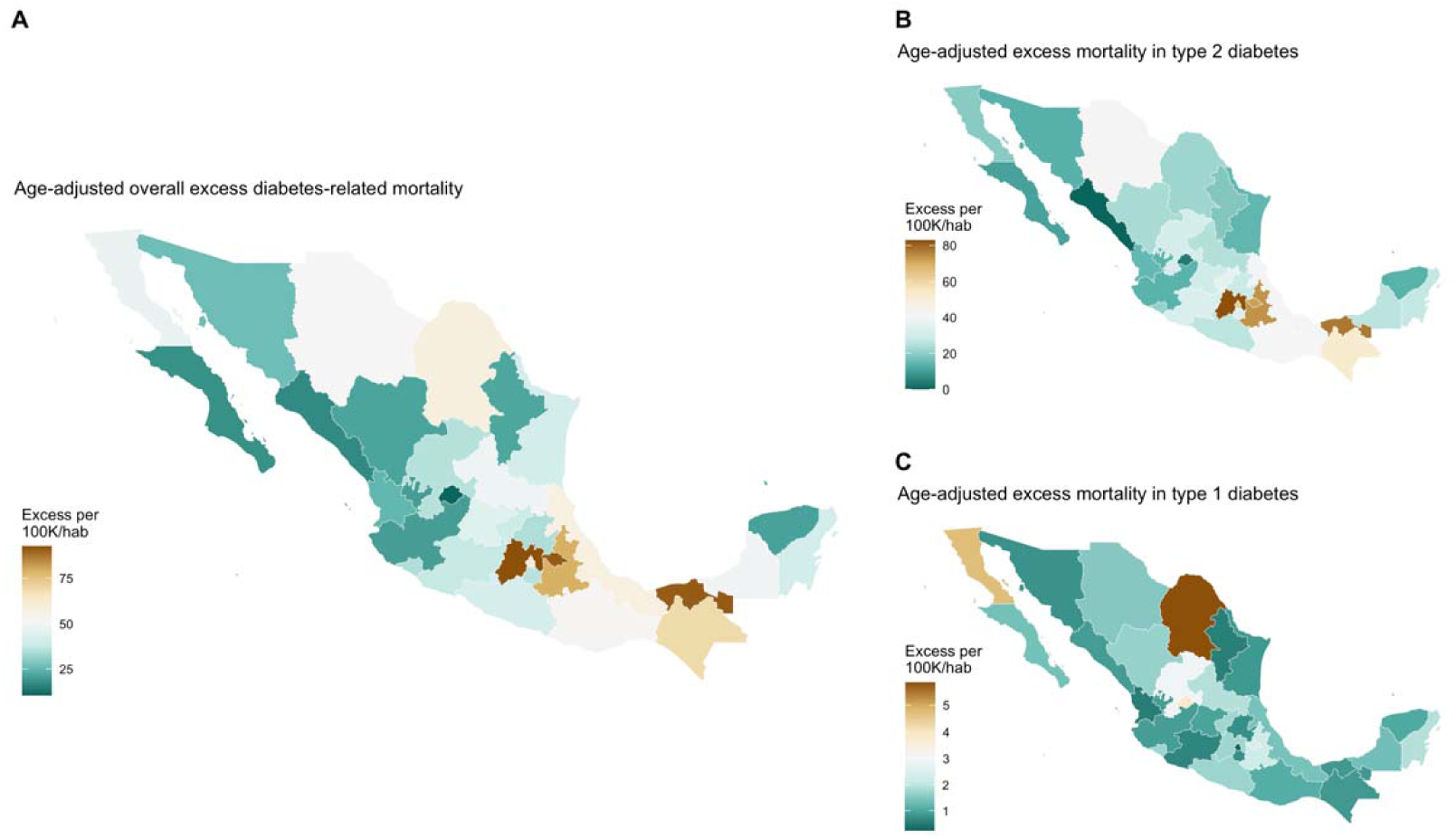
Choropleth maps showing the distribution of estimated age-adjusted diabetes-related excess mortality in Mexico during 2020 compared to 2017-2019, overall (A) and stratified by type 2 (B), and type 1 diabetes (C), according to ICD-10 criteriaDistribution of all evaluated measures was estimated using the quantile method with the *biscale* R package.

**Figure 4.**
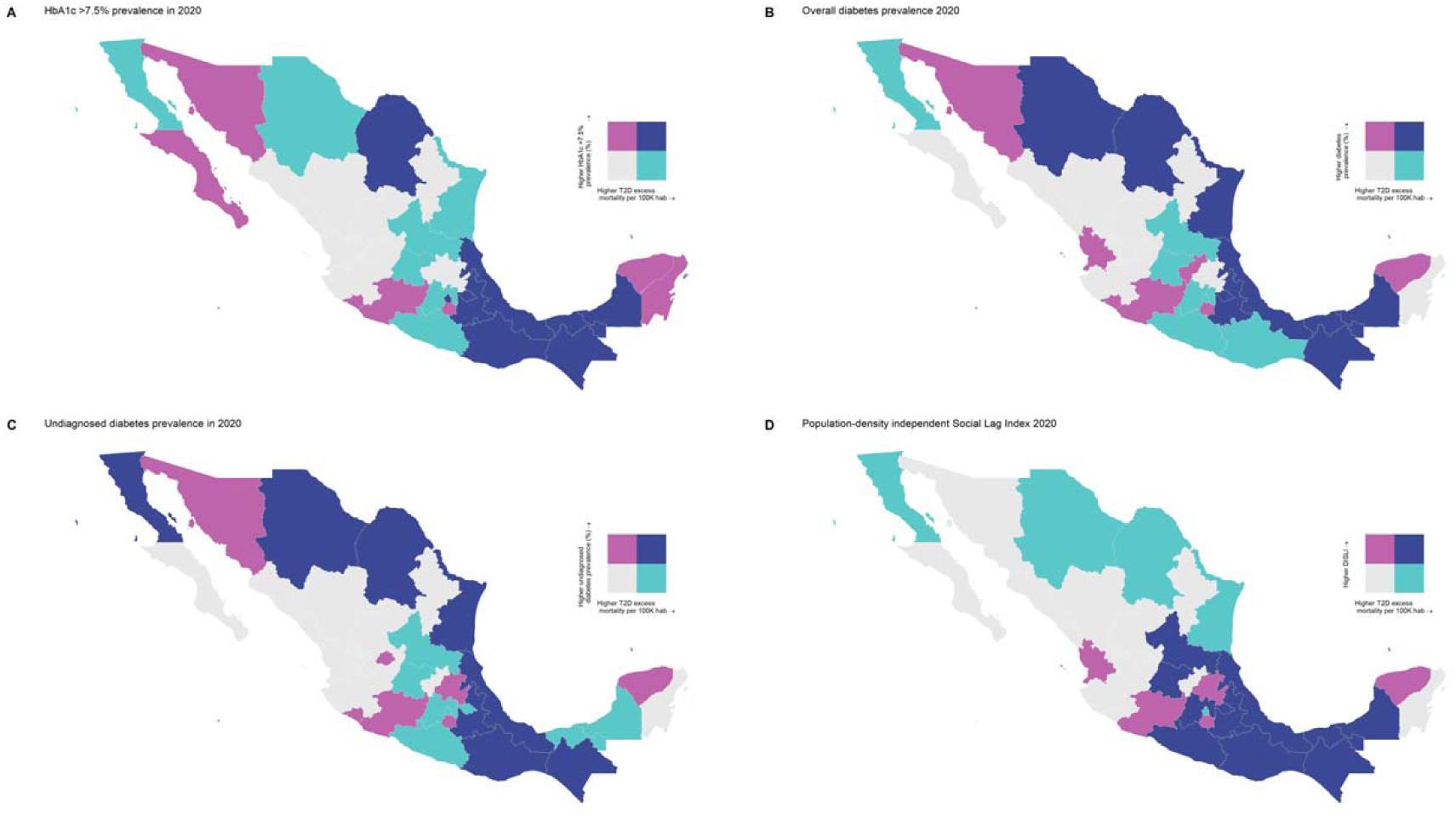
Bivariate choropleth maps showing the geographical distribution of high age-adjusted diabetes-related excess mortality in Mexico with epidemiological indicators related to diabetes care in 2020 including prevalence of HbA1c ≥7.5% (A), diabetes prevalence (B), undiagnosed diabetes prevalence (C), and the 2020 population density-independent social lad index (DISLI, D). Distribution of all evaluated measures was estimated using the quantile method with the *biscale* R package.

We identified a spatial correlation between diabetes-related excess mortality and the DISLI index of marginalization (Lee’s L statistic= 0.326, p<0.001, **Figure 3D-F**). Interestingly, higher DISLI was also correlated with high prevalence of HbA1c levels ≥7.5% (Lee’s L statistic= 0.326, p<0.001). Furthermore, we identified a strong spatial correlation for the out-of-hospital to in-hospital mortality ratio with DISLI (Lee’s L statistic= 0.380, p<0.001) and HbA1c >7.5% prevalence (Lee’s L statistic= 0.247, p=0.047, **Supplementary Figure 7**). We did not identify any relevant spatial correlation between diabetes-related excess mortality and COVID-19 indicators (**Supplementary Figure 8**).

### Association between epidemiologic indicators and diabetes-related excess mortality

To further understand the association between all evaluated epidemiological indicators with age-adjusted diabetes-related excess mortality, we fitted negative binomial regression models. Age-adjusted diabetes-related excess mortality was highest in states with higher DISLI (IRR 1.18, 95%CI 1.01-1.37), higher log-transformed rates of COVID-19 hospitalization (IRR 1.28, 95%CI 1.06-1.55), and higher prevalence of HbA1c ≥7.5% (IRR 1.04, 95%CI 1.01-1.07). The association between higher DISLI and higher diabetes-related excess mortality was particularly pronounced for type 2 diabetes (IRR 1.32, 95%CI 1.08-1.63), which was also associated with higher rates of COVID-19 hospitalization (IRR 1.32, 95%CI 1.01-1.71). Finally, for other diabetes types, age-adjusted diabetes-related excess mortality was associated with higher regional COVID-19 seroprevalence (IRR 1.02, 95%CI 1.01-1.04) and higher overall prevalence of diabetes (IRR 1.04, 95%CI 1.00-1.07) (**Supplementary Table 2**).

## DISCUSSION

In this study of diabetes-related mortality in Mexico between 2017-2020, we identified 452,924 diabetes-related deaths and identified a 41.6% increase in diabetes-related deaths in 2020 (177 per 100,000 inhabitants) compared to the 2017-2019 period (average of 125 per 100,000 inhabitants). Alarmingly, these excess deaths attributable to diabetes occurred disproportionately out-of-hospital, with a 2-fold increase in out-of-hospital vs. in-hospital deaths and an astounding 89.4% increase in out-of-hospital diabetes-related deaths in 2020 compared to 2017-2019. The largest share of diabetes-related deaths in 2020 was attributable to type 2 diabetes (130 deaths per 100,000 habitants), an increase of 46.7% compared to the 2017-2019 period. Diabetes-related excess mortality was also observed for type 1 diabetes by 53.5%. These findings highlight the dramatic increase in diabetes-related mortality that occurred in excess in Mexico in 2020 compared to prior years and underscore the profound impact that the COVID-19 pandemic had on the health system and on access to care for people with diabetes. Urgent policies to strengthen systems of care for people living with diabetes are needed.

We also report a substantial increase in several diabetes-related emergencies and complications as contributing causes of death in 2020 compared to 2017-2019. Notably, HHS and ketoacidosis increased by more than 2-fold in 2020, with these increases being more pronounced in the out-of-hospital setting. This is notable because many hyperglycemic emergencies, especially those related to insulin omission, are preventable; the majority are highly treatable, with mortality rates having dropped substantially with access to inpatient care^19,20^. Consequently, the very high increase in hyperglycemic emergencies can be viewed as an indicator of a profound failure of outpatient access to care and in-patient provision of care since mortality in hyperglycemic emergencies occurred out-of-hospital. Other diabetes-related microvascular complications as contributing causes of death also increased in 2020 compared to prior years, particularly renal, ophthalmic, and circulatory complications. Overall, these findings suggest that interruptions in diabetes care in the setting of the COVID-19 pandemic may have impacted both the acute care of life-threatening diabetes complications, such as HHS and ketoacidosis, as well as the management of chronic complications, such as the need for renal replacement therapy.

Regarding the regional geographical distribution of diabetes-related excess deaths, we identified higher rates of diabetes-related excess mortality in the central and southeast regions of Mexico. These excess deaths were associated with higher socio-demographic inequalities as proxied by DISLI, higher rates of inadequate glycemic control, and increased rates of COVID-19 hospitalizations in 2020. Notably, these regions also had an increase in out-of-hospital and a decrease in in-hospital diabetes-related mortality. Our results are consistent with prior evidence that suggests that the COVID-19 pandemic interfered with routine diabetes care in the region, which intersected with the already high levels of poor glycemic control and social inequalities in Mexico^8,10,11,15^. Since increasing rates of infections and hospital saturation were observed in later COVID-19 waves, a sustained increase in diabetes-related excess mortality is expected. This situation could also lead to a growing burden in diabetes care, with increased likelihood of chronic diabetes complications as a consequence of worsening of glycemic control in Mexico in coming years^7^.

Finally, we identified substantial geographical disparities in the distribution of diabetes-related excess mortality across Mexican States. We identified a cluster of significantly higher diabetes-related excess mortality and large increases in the ratio of out-of-hospital to in-hospital mortality in the southeast region of Mexico; notably, this region displayed a spatial correlation between excess diabetes-related out-of-hospital deaths and both DISLI and increased rates of inadequate glycemic control, indicating that this region experienced the highest impact of socio-demographic inequalities in overall diabetes care. Previous data showed that the impact of diabetes on COVID-19 mortality was largest in the south of Mexico. These findings are particularly important when considering that the southern region of Mexico faces increased socio-economic inequalities, including a limited healthcare infrastructure, which may have impacted quality and access to care for chronic diseases, including diabetes^21^. Our results indicate that this region may be particularly susceptible to hospital saturation during COVID-19 waves and its existing social inequalities as proxied by the DISLI may have a strong association with glycemic control and diabetes-related excess mortality. Special efforts should be sought by Mexican public health authorities to reduce health inequalities within this region, and to strengthen care for diabetes and other cardio-metabolic diseases.

Mechanisms underlying the impact of socio-demographic inequalities on diabetes care and diabetes-related excess mortality in Mexico are not well understood and require further study. Diabetes in Mexico is highly heterogeneous, with data showing a large proportion of diabetes cases relating to obesity and glucotoxicity-mediated decreased β-cell function, but also by socio-demographic inequalities, which markedly impact diabetes care for both type 1 and type 2 diabetes^22–25^. The combined influence of these inequalities likely worsens glycemic control and increases morbidity and mortality in patients living with diabetes. These factors, combined with the ongoing COVID-19 pandemic, which increased hospitalization rates and led to hospital saturation in areas with high population density and high social lag, may have contributed to the observed increase in diabetes-related excess mortality in Mexico^26–28^.

Our study has some strengths and limitations. The strengths of our analysis include the use of several nationally representative datasets, which allowed us to gain insight into potential socio-demographic, epidemiologic, and COVID-19-related correlates of diabetes-related excess mortality in Mexico. Second, we standardized all mortality analyses by age, which allowed for adequate comparisons across Mexican regions with a diverse population structure and more precise estimation of diabetes-related excess mortality. Finally, given that the pandemic had a differential impact across Mexico, we explored spatial effects in the influence of all evaluated epidemiological indicators; this allowed us to understand the regional impact of diabetes-related excess mortality to better inform public policy. We also acknowledge the following limitations which should prompt caution in the interpretation of our results. By using state-level variables, we were unable to perform inferences for all identified associations at individual or even local levels; this is particularly relevant for socio-demographic inequalities, indicators of glycemic control, and COVID-19 seroprevalence which may have significant heterogeneity within Mexican states at both the municipal, local, and individual level. Finally, since ascertainment of COVID-19 cases in Mexico has been insufficient^17^, many SARS-CoV-2 infections could have been undetected and some of these may have led to diabetes-related complications such as diabetic ketoacidosis or HHS^29,9^. Therefore, we cannot rule out that a portion of excess mortality formally ascribed to diabetes could have been in fact in part attributable to COVID-19, despite the fact that ascertainment by INEGI is generally robust^30^.

In conclusion, we report a 41.6% increase in diabetes-related mortality in Mexico in 2020 after excluding COVID-related death, compared to the 2017-2019 period, with excess deaths largely attributed to type 2 diabetes and disproportionately occurring out-of-hospital. Diabetes-related complications as contributing causes of death also increased dramatically in 2020, particularly HHS and diabetic ketoacidosis. Diabetes-related excess mortality displayed significant geographic heterogeneity, with clusters of high excess mortality in the Southeast and Gulf regions of Mexico. Socio-demographic inequalities as proxied by the DISLI, a high prevalence of HbA1c ≥7.5%, and higher rates of COVID-19 hospitalization were associated with increased diabetes-related excess mortality. Our results could help inform policies to reduce diabetes-related mortality in Mexico, mitigate the impact of the COVID-19 pandemic on people with diabetes, and reduce socio-demographic inequalities, particularly in areas of Mexico with high marginalization. More broadly, hyperglycemic emergencies and conditions in patients with diabetes and chronic kidney disease exemplify conditions that are readily treated in a functioning healthcare system but can cause substantial short-term increases in mortality when inpatient hospital access is restricted. Given that the COVID-19 pandemic is still ongoing, policies are urgently needed to prevent further sustained increases in diabetes-related mortality.

## Data Availability

All code, datasets and materials are available for reproducibility of results at https://github.com/oyaxbell/diabetes_excess/

## ACKNOWLEDGMENTS

This project was registered and approved by the Research Committee at Instituto Nacional de Geriatría, project number DI-PI-006/2020. NEAV and CAFM are enrolled at the PECEM Program of the Faculty of Medicine at UNAM. NEAV is supported by CONACyT.

## AUTHOR CONTRIBUTIONS

Research idea and study design: OYBC, NEAV, CAFM, LFC, RCL, JMG, JAS; data acquisition: OYBC, NEAV, CAFM, LFC; analysis/interpretation: OYBC, NEAV, CAFM, LFC; statistical analysis: CAFM, AMS, OYBC; manuscript drafting: OYBC, NEAV, CAFM, LFC, AVV, MRBA, DRG, AESL, RCL, DJW, JMG, JAS; supervision or mentorship: OYBC. Each author contributed important intellectual content during manuscript drafting or revision and accepts accountability for the overall work by ensuring that questions pertaining to the accuracy or integrity of any portion of the work are appropriately investigated and resolved.

