## Supplementary Material for "Diabetes-related excess mortality in Mexico: a comparative analysis of national death registries between 2017-2019 and 2020"

**SUPPLEMENTARY METHODS**

*Mexican regions*

For all mortality analyses, cases were aggregated at the state level for each of the 32 states in Mexico. For analyses in Supplementary Table 1 rates were analyzed at 9 larger regions defined by ENSANUT COVID 2020 because of their geographical proximity and similar population density. Regions were classified as Northern Pacific (including Baja California, Baja California Sur, Nayarit, Sinaloa, Sonora; representing 9% of the population), Border region (Chihuahua, Coahuila, Nuevo León, Tamaulipas, 12%), Central Pacific (Colima, Jalisco, Michoacán, 11%), Northern Central (Aguascalientes, Durango, Guanajuato, Querétaro, San Luís Potosí, Zacatecas, 13%), Central region, (Hidalgo, Tlaxcala, Veracruz, 10%), Mexico City (8%), Mexico State (14%), Southern Pacific (Guerrero, Morelos, Oaxaca, Puebla, 13%) and Peninsula region (Campeche, Chiapas, Quintana Roo, Tabasco, Yucatán, 10%).

*ENSANUT COVID 2020*

To evaluate potential correlated excess mortality related to the epidemiology of diabetes in Mexico, we extracted these estimates from the Mexican National Health and Nutrition Survey COVID carried out in 2020 (ENSANUT COVID 2020). ENSANUT is a population-based survey which aims to evaluate the health and nutritional status of Mexican adults, and which is representative at a national, regional, and rural/urban level. ENSANUT uses two-stage probabilistic cluster stratified sampling based on households and individuals and has been conducted in 2006, 2012, 2016, 2018 and 2020. ENSANUT COVID 2020 was collected from August to November 2020, recruited 24,726 adults ≥20 years, and was used to estimate diabetes-related prevalence and COVID-19 seroprevalence. This survey additionally aimed to investigate health and well-being of Mexican adults during the COVID-19 pandemic. A random subsample of 16,150 adults were selected to undergo biochemical testing to assess for presence of SARS-CoV-2 antibodies, with a 47% participation rate (n=7,573). A second subsample (n=2,373) had an additional biochemical evaluation with serum samples for glycated hemoglobin (HbA1c), fasting glucose, and a fasting lipid profile.

*Density-independent social lag index*

To quantify the impact of sociodemographic inequalities on diabetes-related mortality during 2020 at a state level, we used the 2020 social lag index (SLI), a composite assessment of the degree of healthcare access, economic well-being, and access to basic services in Mexico^1^. Population density was calculated as proposed by INEGI for all states. Because we intended to evaluate inequalities independent of population density, we used residuals of linearly regressed population density onto SLI values to approximate a Density-independent SLI (DISLI), as previously validated for Mexico City^2^.

*Diabetes definitions*

Diabetes was defined by self-report among individuals who answered “yes” to the question “Has a doctor ever told you that you have diabetes or high blood sugar?” or by either a fasting blood glucose level of ≥126 mg/dL or a hemoglobin A1c (HbA1c) ≥6.5%. Individuals who met the biochemical definition of diabetes but who responded “no” to a prior diagnosis of diabetes were categorized as having undiagnosed diabetes. Adequate glycemic control was defined based on an HbA1c level of ≥7.5%^3^. The prevalence of diabetes according to these definitions was estimated using individual-level data from the ENSANUT COVID 2020 at both region and state levels.

COVID-19 seroprevalence

Seropositivity to SARS-CoV-2 was evaluated using a Elecsys detection assay for IgG against the N-protein (SARS-CoV-2 Nucleocapsid, Roche) validated by the Institute for Epidemiological and Diagnostic Reference using data from ENSANUT COVID 2020 at both region and state levels. Samples were considered positive if quantification was ≥1.0 U/ml^4^.

*COVID-19 incidence, hospitalization, and mortality*

State-wide incidence, hospitalization, and deaths attributable to COVID-19 in Mexico were evaluated using data from the General Directorate of Epidemiology of the Mexican Ministry of Health, which is an open-source dataset that provides daily updated information of suspected COVID-19 cases. COVID-19 caseshave been confirmed with a positive RT-PCR or rapid antigen test for SARS-CoV-2 andwere certified by the National Institute for Diagnosis and Epidemiological Referral until December 31^st^, 2020^5–7^. Estimates were obtained by COVID-19 cases with comorbid diabetes. All metrics were weighted to their respective population by state to reflect rates per 100,000 inhabitants using data from National Population Council of Mexico (CONAPO).

**SUPPLEMENTARY RESULTS**

*Geographic variability in age-adjusted diabetes-related excess mortality in Mexico*

Overall, age-adjusted diabetes-related mortality was highest in Mexico State, Tabasco, and Tlaxcala, with a cluster of high diabetes-related excess mortality located in the Southeast of Mexico. When stratified by diabetes type, type 2 diabetes-related excess mortality mirrored that of overall diabetes-related excess mortality, while type 1 diabetes-related excess mortality was highest in the northern states of Baja California, Chihuahua, and Aguascalientes (**Figures 3A-C**). Mortality for other diabetes types was highest in the northern states of Baja California, Coahuila, and Tamaulipas. In bivariate analyses, we identified that a cluster of states with high age-adjusted diabetes-related excess mortality and high prevalence of HbA1c levels ≥7.5% was observed in the Southeast region of Mexico and in the state of Chihuahua, with a similar trend observed for diabetes prevalence (**Figures 3D-F**).

**SUPPLEMENTARY FIGURES**

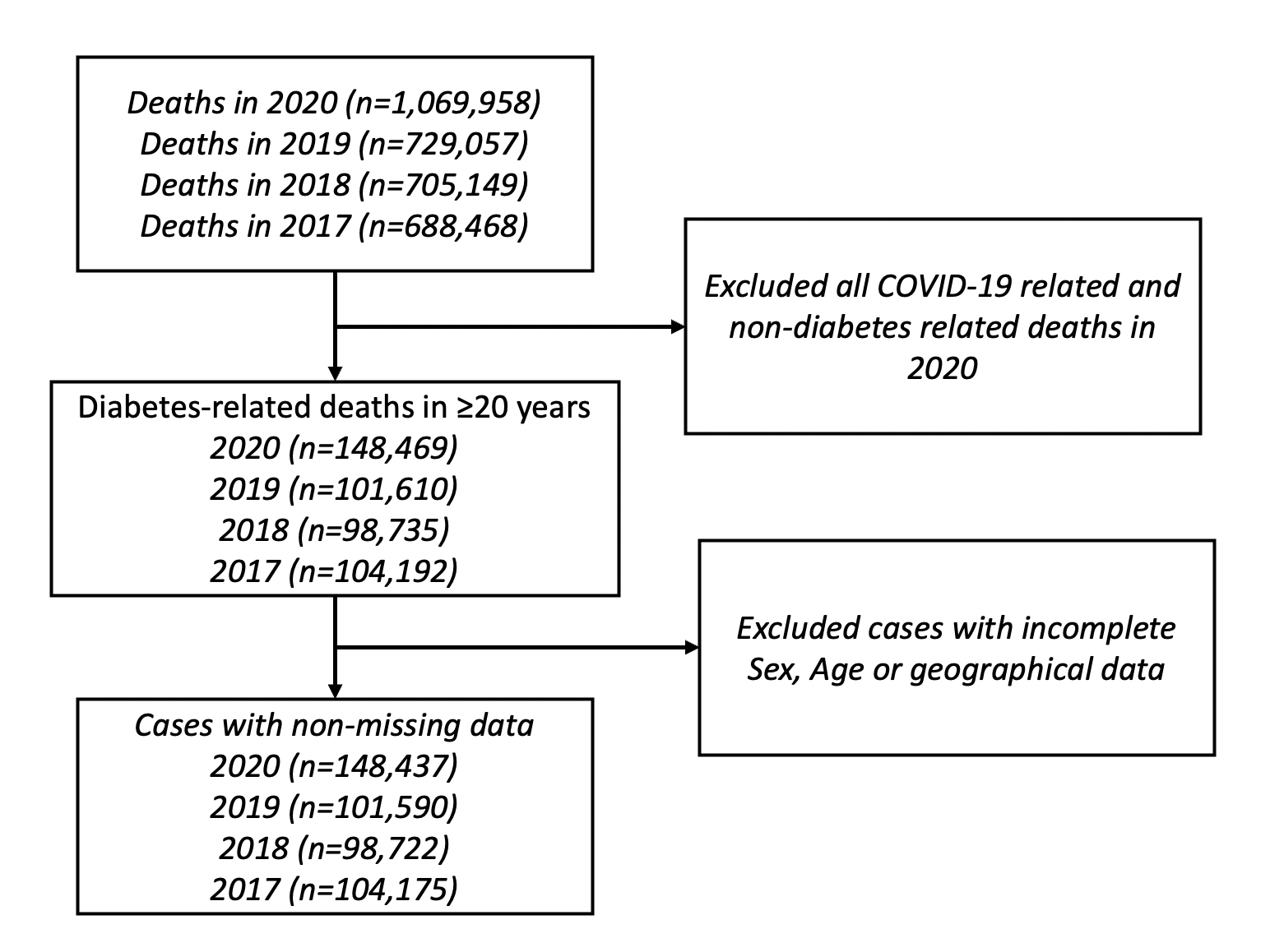

**Supplementary Figure 1.** Flowchart diagram of data selection for the study in the mortality dataset from 2017-2020.

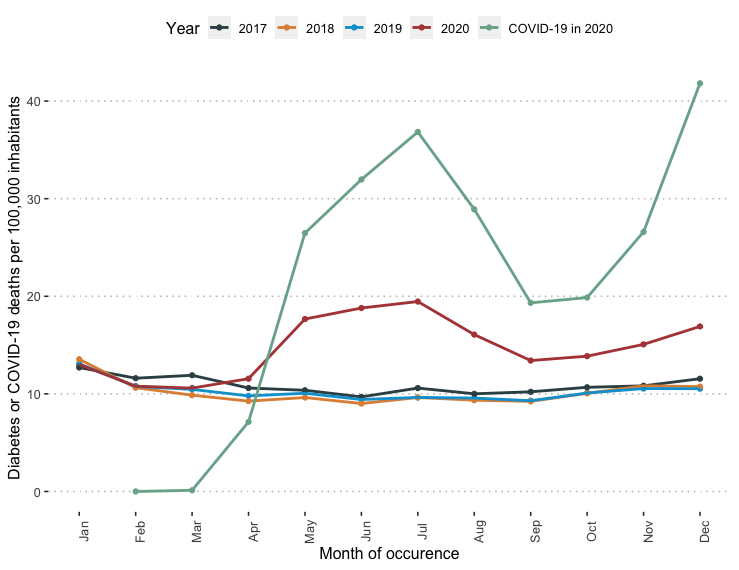

**Supplementary Figure 2.** Diabetes-related mortality in the 2017-2020 period (ICD-10 codes E10-E14) compared to COVID-19 related deaths in 2020 (ICD-10 codes U07.1 U07.2).

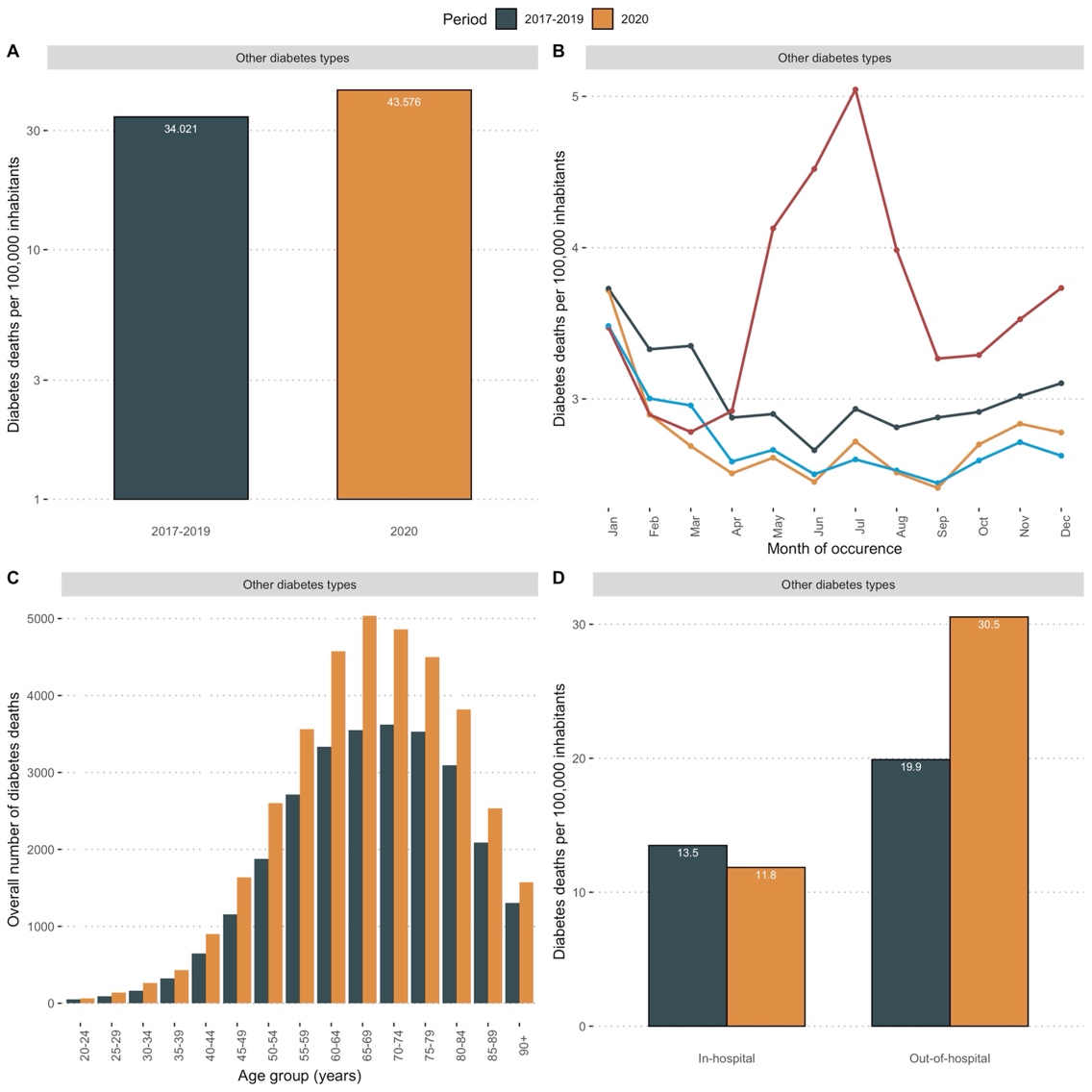

**Supplementary Figure 3.** Diabetes-related mortality for other diabetes types during the 2017-2019 period compared to 2020 (A) and stratified by month (B), age group by 5-year increments (C) and in-hospital vs. out-of-hospital setting (D).

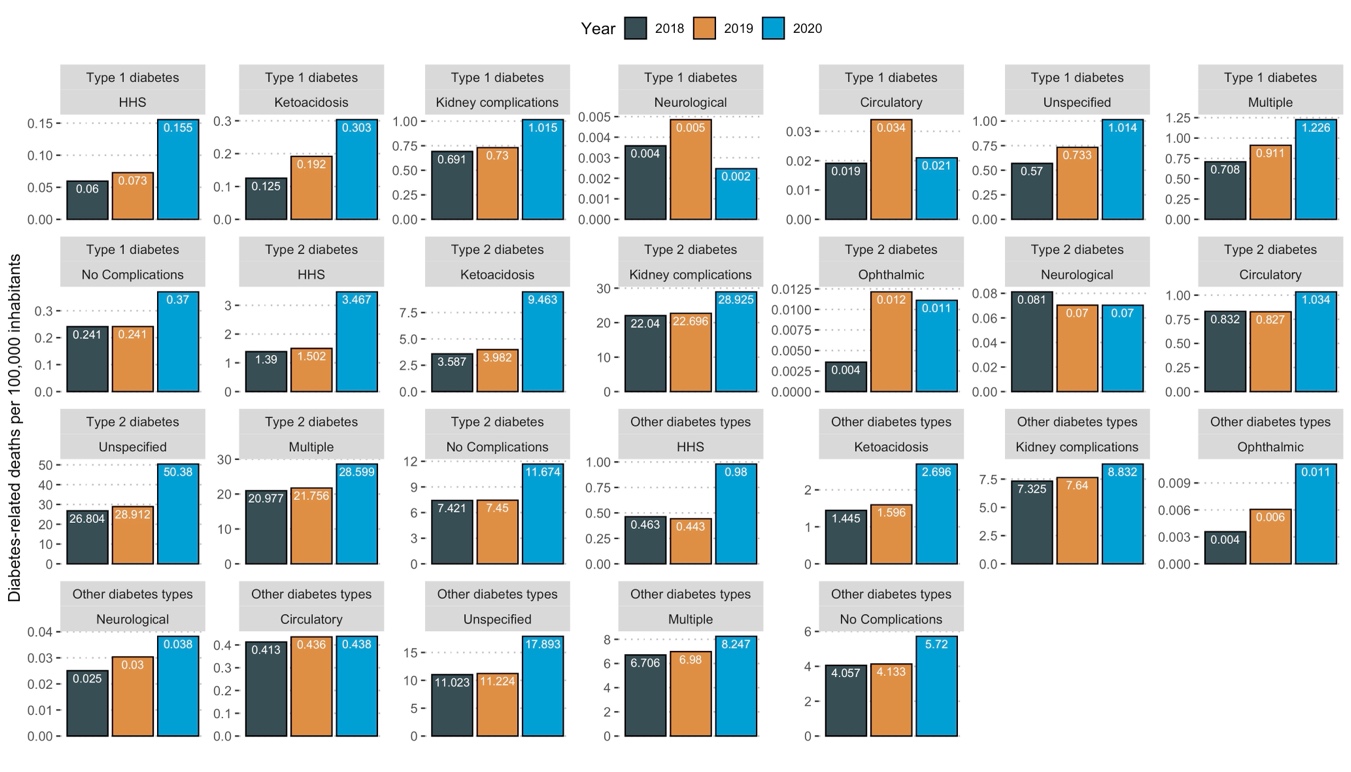

**Supplementary Figure 4.** Mortality rates for diabetes-related emergencies and complications as contributing causes of death in the 2018-2020 period, stratified by diabetes type, standardized to 100,000 inhabitants. Deaths were categorized using ICD-10 codes.

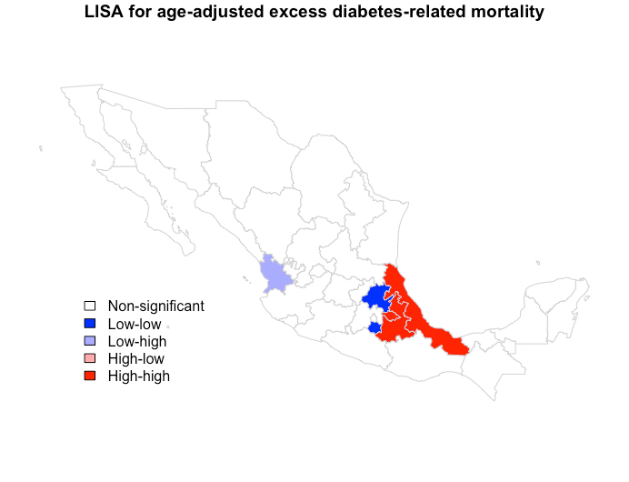

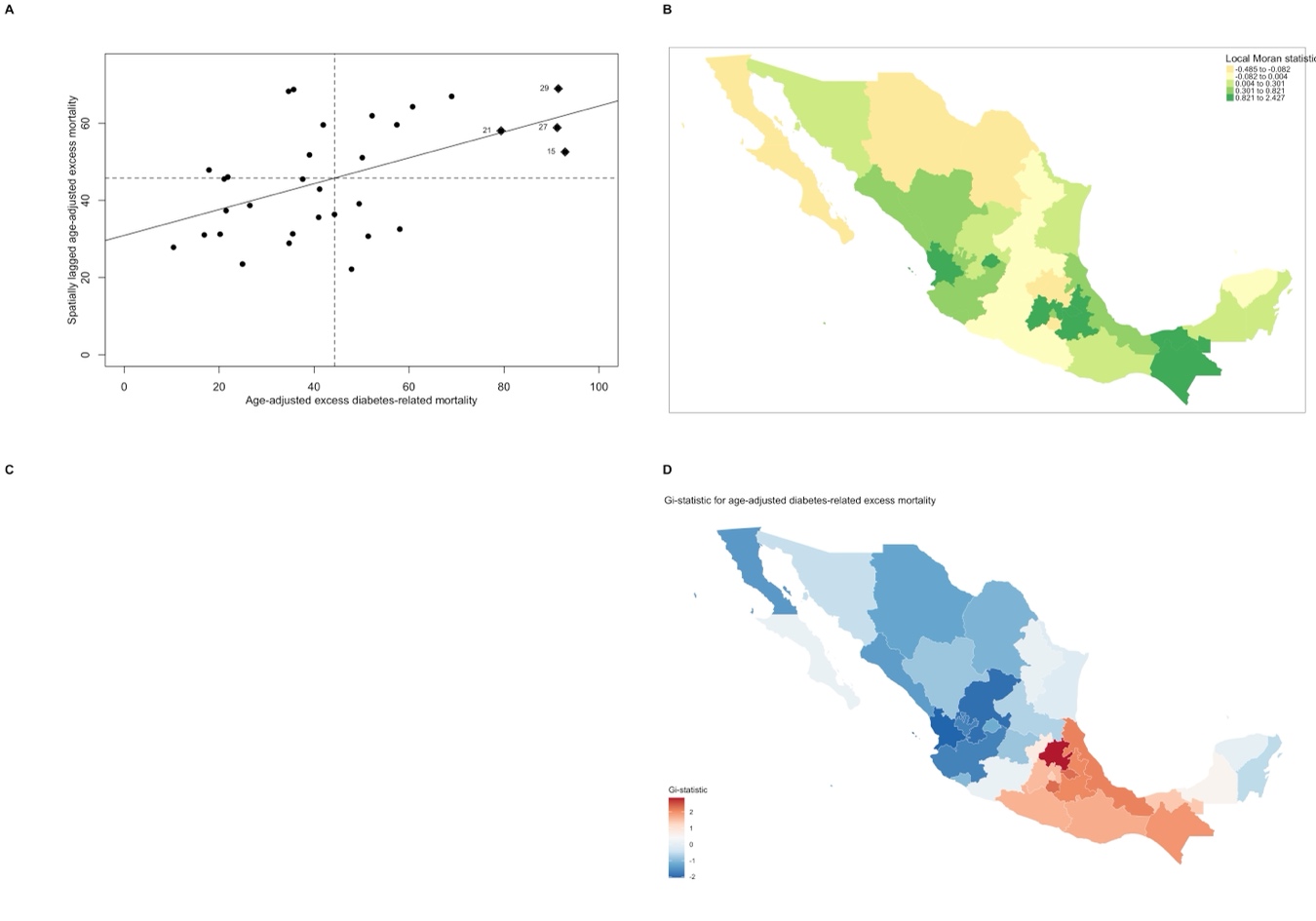

**Supplementary Figure 5.** Spatial autocorrelation of age-adjusted excess diabetes-related mortality in Mexico using Moran’s I statistic (A). Figure also shows the distribution of Local Moran Statistic (B) and the local indicators of spatial autocorrelation (LISA) using Moran’s I statistic for age-adjusted diabetes-related excess mortality. Panel C shows a cluster of states high diabetes-related excess mortality surrounded by similar states in the Gulf of Mexico Region (C). Panel D shows the distribution of the Getis-Ordi Gi statistic, representing hot spots of diabetes-related excess mortality primarily located in southern Mexico and the Gulf region (D).

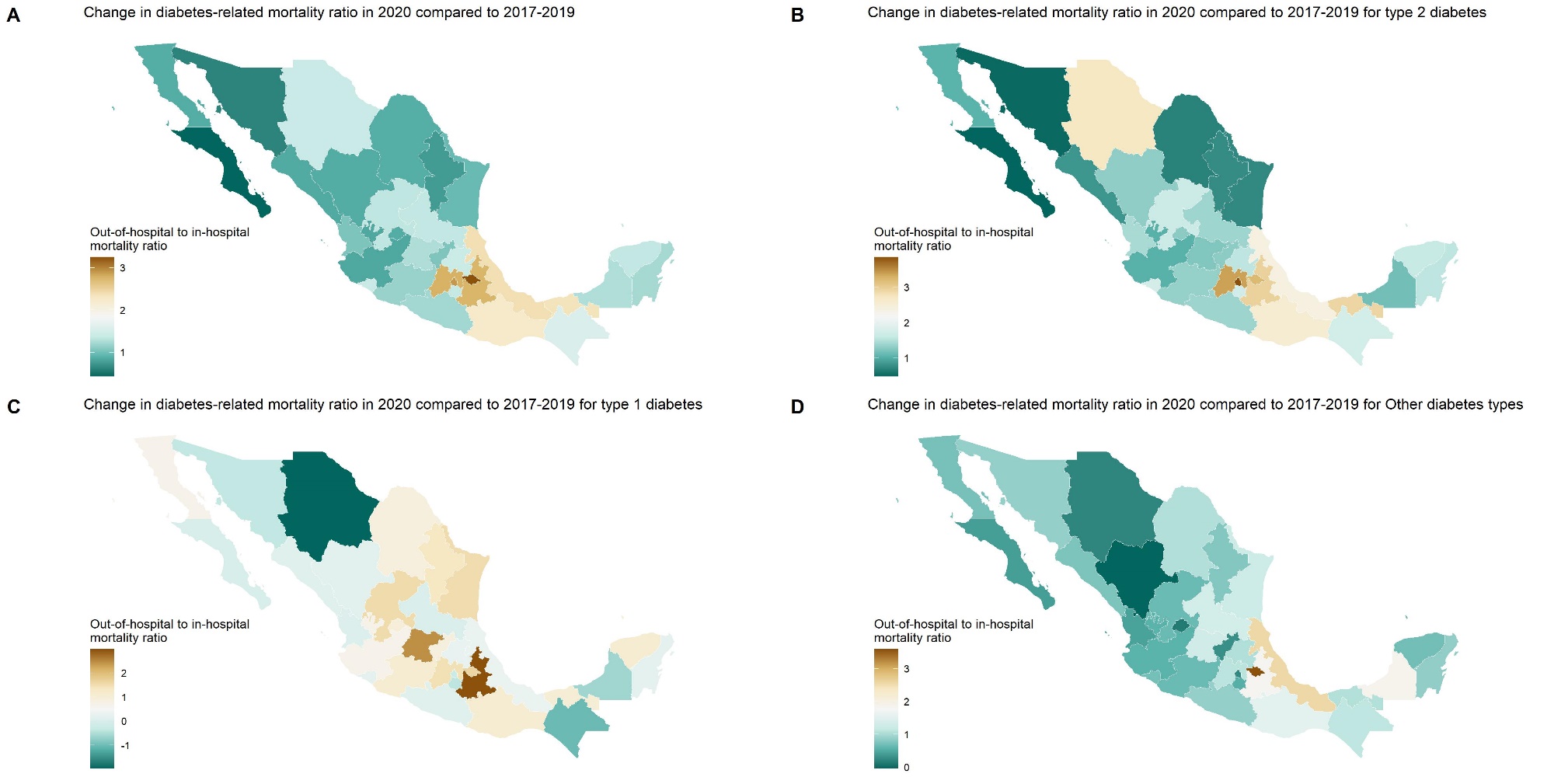

**Supplementary** **Figure 6.** Chloropleth maps showing the geographical distribution of the difference in out-of-hospital to in-hospital death ratio in 2020 compared to the average of 2017-2019 for overall diabetes-related mortality (A), and stratified for type 2 (B), type 1(C) and other types of diabetes (D), codified using ICD-10 causes of death registered by INEGI and the Mexican Ministry of Health.

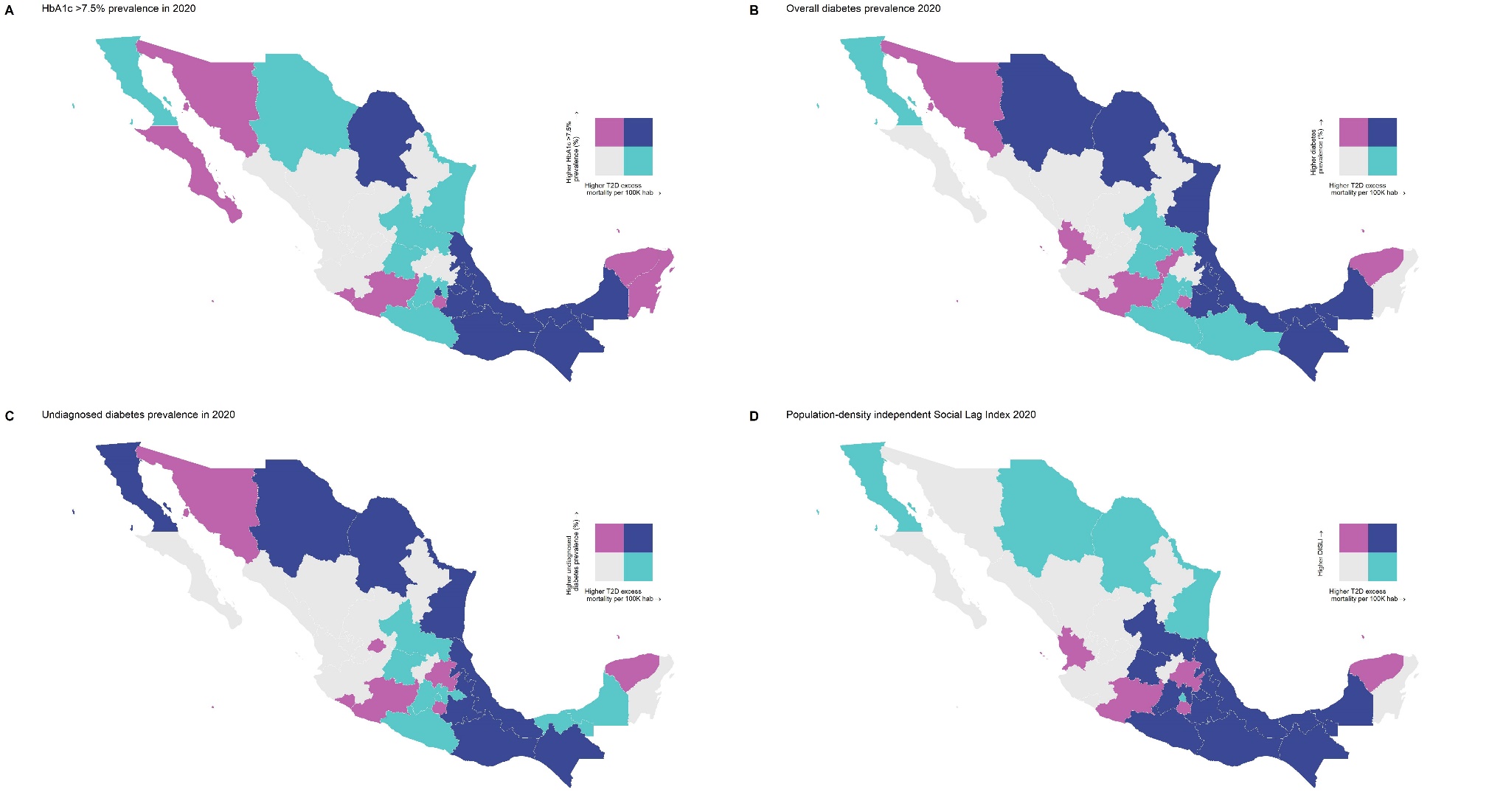

**Supplementary** **Figure 7.** Bivariate chloropleth maps showing the geographical distribution of high out-of-hospital to in-hospital diabetes-related mortality in Mexico with epidemiological indicators related to diabetes care including HbA1c >7.5% prevalence (A), overall diabetes prevalence (B), undiagnosed diabetes prevalence (C), and the population-density independent social lag index (DISLI, D). Distribution of all evaluated measures was estimated using the quantile method with the *biscale* R package.

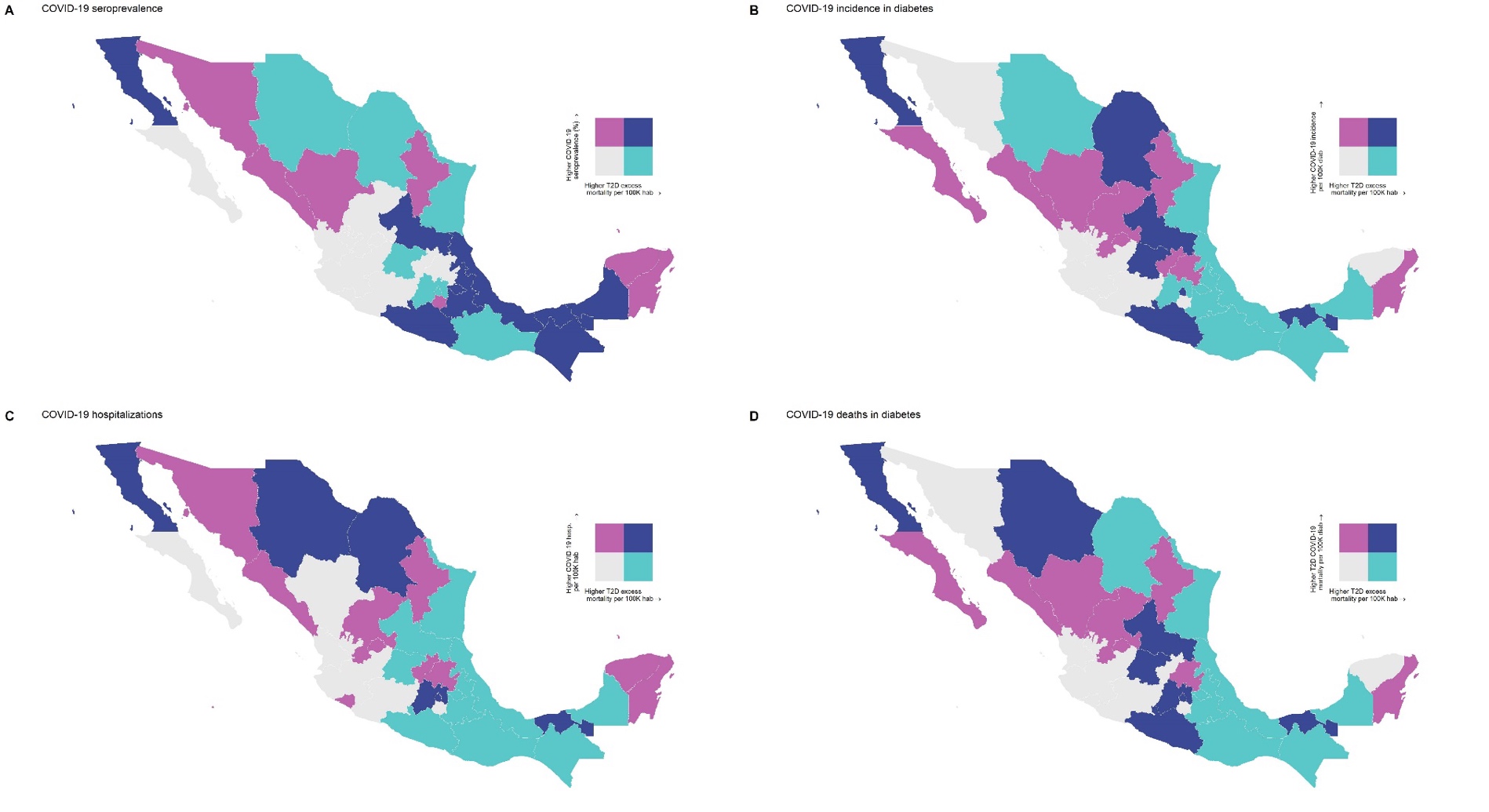

**Supplementary** **Figure 8.** Bivariate chloropleth maps showing the geographical distribution of high diabetes-related excess mortality in Mexico with epidemiological indicators related to COVID-19 including COVID-19 seroprevalence (A), COVID-19 incidence in diabetes (B), COVID-19 hospitalizations (C), and COVID-19 deaths in diabetes (D). Distribution of all evaluated measures was estimated using the quantile method with the *biscale* R package.

**SUPPLEMENTARY TABLES**

**Supplementary Table 1.** Age-adjusted excess mortality rate per 100,000 habitants in nine Mexican regions during 2020, along with prevalence of diabetes, undiagnosed diabetes, HbA1c ≥7.5% and COVID-19 seroprevalence as estimated using ENSANUT COVID 2020.

| Region | Diabetes mortality 2017-2019 | Diabetes mortality 2020 | Age-adjusted excess mort. rate | Diabetes prev. (%) | Undiagnosed Diabetes (%) | HbA1c ≥7.5% (%) | COVID-19 seroprev. (%) |
| --- | --- | --- | --- | --- | --- | --- | --- |
| Mexico City | 118.05 | 178.79 | 60.74 | 14.53 | 2.94 | 7.15 | 18.40 |
| Center | 137.39 | 192.68 | 55.29 | 22.19 | 9.56 | 13.45 | 23.85 |
| Center-North | 122.90 | 161.14 | 38.24 | 12.33 | 2.50 | 3.81 | 17.93 |
| Mexico State | 143.74 | 236.60 | 92.86 | 13.24 | 2.58 | 6.59 | 21.99 |
| Frontier | 105.59 | 145.67 | 40.08 | 17.05 | 4.89 | 6.82 | 19.73 |
| South pacific | 140.49 | 198.69 | 58.20 | 15.91 | 4.84 | 7.27 | 22.76 |
| Center Pacific | 117.49 | 144.78 | 27.29 | 14.76 | 4.48 | 6.27 | 18.26 |
| Northern Pacific | 91.66 | 121.24 | 29.58 | 14.43 | 4.62 | 10.40 | 28.85 |
| Península | 135.41 | 194.42 | 59.01 | 16.77 | 6.96 | 11.40 | 39.74 |

**Abbreviations:** Prev., prevalence; seroprev., seroprevalence.

| Model | Parameter | IRR (95%CI) | p-value |
| --- | --- | --- | --- |
| Overall  R^2^=0.30 | Intercept | 3.78 (0.66-21.51) | 0.13 |
|  | DISLI | 1.18 (1.01-1.37) | 0.03 |
|  | COVID-19 hospitalization | 1.28 (1.06-1.55) | 0.01 |
|  | Prevalence of HbA1c ≥7.5% | 1.04 (1.01-1.07) | 0.03 |
| Type 2 diabetes  R^2^=0.23 | Intercept | 2.88 (0.29-29.03) | 0.37 |
|  | DISLI | 1.32 (1.08-1.63) | 0.01 |
|  | COVID-19 hospitalization | 1.32 (1.01-1.71) | 0.04 |
| Other diabetes types  R^2^=0.20 | Intercept | 4.90 (2.33-10.29) | <0.01 |
|  | COVID-19 seroprevalence | 1.02 (1.01-1.04) | 0.02 |
|  | Diabetes prevalence | 1.04 (1.01-1.07) | 0.03 |
| Type 1 diabetes  R^2^=0.07 | Intercept | 1.71 (0.80-3.62) | 0.16 |
|  | DISLI | 1.00 (0.97-1.03) | 0.84 |
|  | Diabetes prevalence | 0.80 (0.57-1.12) | 0.20 |

**Supplementary Table 2.** Negative binomial regression models to assess association of epidemiological indicators of age-adjusted diabetes-related excess mortality stratified by diabetes type. **Abbreviations:** DISLI, Density-independent social lag index; IRR, Incidence Rate Ratio; 95%CI, 95% Confidence interval.
